# Urban Rural Disparities of Determinants of Antenatal Care in Senegal among Women of Reproductive Age: Evidence from 2023 Senegal Demographic Health Survey

**DOI:** 10.1101/2024.10.12.24315378

**Authors:** Mushfiqur Rahman Khan Majlish, Sultanul Arafean Tawhid, Gulam Kibria

## Abstract

**Background:** Antenatal care (ANC) is critical for ensuring positive pregnancy outcomes. Despite global efforts, ANC utilization remains suboptimal in many developing countries, including Senegal. This study aims to identify and distinguish between the key socio-demographic and economic factors influencing ANC utilization among urban and rural women in Senegal.

**Methods:** The analysis was conducted on 5107 women who had at least one birth in the last five years preceding the survey. Data were obtained from the 2023 Senegal Demographic and Health Survey (DHS). ANC utilization, defined as receiving four or more ANC visits during pregnancy, was the outcome variable. Independent variables included age, region, education, wealth index, total children ever born, pregnancy intention, employment status, media exposure, births in last five years and women autonomy. Bivariate analyses and multiple logistic regression were conducted to identify the significant variables.

**Results:** The final sample included 4,785 women after excluding those with missing data. Higher education level (Adjusted Odds Ratio (AOR) = 3.649), rich wealth index (AOR=1.581), low (AOR=1.792) and high (AOR=1.692) media exposure were significantly associated with higher ANC in urban residences. On the contrary, in rural residences, primary (AOR=1.60), secondary (AOR=2.01) and higher (AOR=5.79) education level, intended pregnancy (AOR=1.33), low (AOR=1.48) and high (OR=1.35) media exposure and women autonomy (AOR=1.29) were significantly associated with higher ANC.

**Conclusions:** This study underscores the influence of different socio-economic and demographic factors on ANC utilization in urban and rural residences in Senegal. Policy interventions should prioritize enhancing education, economic opportunities, media awareness and women’s autonomy, particularly in rural areas, to improve ANC coverage.

## Introduction

Antenatal care (ANC) refers to the healthcare services provided to women during pregnancy by trained health professionals [1]. The key objectives of ANC include the prevention, detection, and management of pregnancy-related complications, ensuring safe delivery, and promoting the well-being of both the mother and the baby. ANC services generally include identifying potential complications during pregnancy, evaluating pregnancy risks, addressing any issues that may occur during the antenatal phase, administering treatments to enhance pregnancy outcomes, offering guidance to expectant mothers, and helping them prepare both physically and mentally for childbirth and parenthood [2]. The World Health Organization (WHO) recommended at least four ANC visits for focused care, if the woman was eligible [3].

Maternal deaths due to pregnancy and childbirth complications have declined from 451,000 in 2000 to 287,000 in 2020, despite significant population growth in regions with high maternal mortality rates. Yet, nearly 800 women continue to die daily from pregnancy-related complications, equating to one death every two minutes [4]. In West and Central Africa, maternal mortality decreased from 890 per 100,000 live births in 2000 to 724 in 2020. In sub-Saharan Africa, the rate dropped from 802 to 536, and in South Asia, it fell from 417 to 138 during the same period [5]. Reducing maternal mortality is a critical component of achieving the Sustainable Development Goals 3 (SDGs) which focuses on good health and well-being [6].

The factors influencing ANC utilization in African countries are diverse, ranging from socioeconomic status and education levels to geographic location and cultural practices. Barriers to accessing ANC include financial limitations, inadequate healthcare infrastructure, and a lack of awareness about the importance of regular ANC visits. For example, poor infrastructure and insufficient readiness of health facilities negatively impact ANC quality in countries like Ethiopia, Kenya, and South Africa [4]. Additionally, higher education levels and media exposure have been associated with increased ANC uptake, as seen in studies from East African countries [7]. Moreover, it was also revealed that distance to health facility and maternal autonomy were significantly associated with ANC in Kenya [8]. These findings underscore the necessity for tailored interventions to enhance ANC utilization across Africa.

While ANC is recognized for its critical role in improving maternal and neonatal health outcomes, research specifically focusing on ANC in Senegal remains limited. Although several studies have investigated the effectiveness of ANC in African countries such as Ethiopia, Kenya, and Nigeria, no dedicated research paper has been published on Senegal’s ANC practices. This research gap underscores the need for focused studies that address the distinct healthcare challenges faced by pregnant women in Senegal. For instance, studies had explored ANC quality in low- and middle-income countries but did not specifically address Senegal [4]. Similarly, broader reviews by organizations like the WHO examine ANC’s impact on maternal mortality globally without providing Senegal-specific data [9]. This gap highlights the need for more research to guide healthcare policies and practices in Senegal.

## Materials and Methods

### Data Source and Population

This study made use of secondary data from the 2023 Senegal Demographic and Health Survey, which was carried out from February to August of that year and was nationally representative. There were two stages in the stratified sample process that the Demographic Health Survey Authority used. 14 regions—Dakar, Ziguinchor, Diourbel, Saint-Louis, Tambacounda, Kaolack, Thies, Louga, Fatick, Kolda, Matam, Kaffrine, Kedougou, and Sedhiou—were used to gather the data. A vast array of information was gathered from 8423 homes, 16583 female respondents who were between the ages of 15 and 49, and 6321 male respondents who were between the ages of 15 and 59. Among other important public health concerns, these included adult and pediatric morbidity and mortality, knowledge of and attitudes toward Human immunodeficiency viruses (HIV), fertility and fertility desires, marriage, and different elements of reproductive health. The study was conducted on those women from urban and rural residence who had minimum one birth in last five years prior to the survey and the number of respondents was 5107.

### Study Variables

The dependent variable was whether the respondent received ANC at the time of pregnancy which has 2 types of responses: either ANC received or did not receive. According to WHO, a respondent should receive minimum four ANC visits during her pregnancy [3]. Hence whether the respondent had received ANC or not was recoded as:

1. No (0) if the respondent had less than four antenatal care visits.
2. Yes (1) if the respondent had minimum of four antenatal care visits.

The explanatory variables that were used in this study are respondent’s age group (15-19, 20-24, 25-29, 30-34, 35-39, 40-44 and 45-40), region (Sedhiou, Dakar, Ziguinchor, Diourbel, Saint-Louis, Tambacounda, Kaolack, Thiès, Louga, Fatick, Kolda, Matam, Kaffrine, Kédougou), respondent’s education level (no education, primary education, secondary, higher), wealth index (poor, middle, rich), total children born (less than 2, 2 to 4, more than 4), pregnancy intention (intended, unintended), employment status (not working, working), media exposure (no exposure, low exposure, high exposure), births in last five years (one birth, two births, more than two births) women autonomy (no, yes).

### Data Processing and Analysis

Several variables had small number of missing values (<5%) and those observations were deleted for the analysis purpose. The final dataset had reduced observation of 4785. The study focuses on identifying different determinants of urban and rural respondents that are responsible for receiving ANC visits. Therefore, two different datasets were extracted containing urban and rural respondent’s information separately from the original dataset. The number of respondents on urban and rural were 1639 and 3146. Simple frequency distribution was conducted on the original dataset to understand the background characteristics. Utilizing a chi-square test and basic cross-tabulation, the bivariate relationship between the independent and dependent variables was examined in urban and rural datasets separately. Finally, multiple logistic regression was conducted to find the association between ANC utilization and independent variables. The STATA version 14.2 was utilized to conduct the entire analysis.

## Results

### Background Characteristics

Table 1 provides an in-depth analysis of several demographic and socio-economic factors affecting the population under investigation. The data reveals diverse educational backgrounds among respondents, with a significant 57.7% having no formal education, highlighting a major educational gap. About 18.7% have completed primary education, 21.1% have secondary education, and only 2.4% have higher education, indicating limited access to advanced education. Geographic distribution varies, with Sedhiou and Diourbel making up 7.9% and 7.5% of the sample, respectively, and Kaffrine having the largest share at 11.1%. Ziguinchor represents just 3.5%, suggesting differences in population density and socio-economic conditions across regions. Most respondents (65.7%) live in rural areas, compared to 34.3% in urban areas, highlighting a predominant rural demographic. Regarding wealth, 57.8% of respondents are classified as poor, 18.8% as middle, and 23.4% as rich, reflecting significant economic challenges. 49.5% of respondents had 2-4 children, 29.0% of respondents had more than 4, and 21.5% of respondents had fewer than 2 children. 53.6% of respondents had one birth in the last five years, 41.7% had two births, and only 4.7% reported more than two births.

**Table 1:** Background characteristics of the study participants.

| Variable | Category | Frequency | Percent (%) |
| --- | --- | --- | --- |
| Educational Level | No education | 2763 | 57.7 |
|  | Primary | 896 | 18.7 |
|  | Secondary | 1009 | 21.1 |
|  | Higher | 117 | 2.4 |
| Region | Sedhiou | 377 | 7.9 |
|  | Dakar | 276 | 5.8 |
|  | Ziguinchor | 166 | 3.5 |
|  | Diourbel | 361 | 7.5 |
|  | Saint-Louis | 305 | 6.4 |
|  | Tambacounda | 373 | 7.8 |
|  | Kaolack | 336 | 7.0 |
|  | Thiès | 365 | 7.6 |
|  | Louga | 312 | 6.5 |
|  | Fatick | 389 | 8.1 |
|  | Kolda | 371 | 7.8 |
|  | Matam | 294 | 6.1 |
|  | Kaffrine | 533 | 11.1 |
|  | Kedougou | 327 | 6.8 |
| Residence | Urban | 1639 | 34.3 |
|  | Rural | 3146 | 65.7 |
| Wealth index | Poor | 2767 | 57.8 |
|  | Middle | 900 | 18.8 |
|  | Rich | 1118 | 23.4 |
| Total Children Born | Less than 2 | 1027 | 21.5 |
|  | 2 to 4 | 2368 | 49.5 |
|  | More than 4 | 1390 | 29.0 |
| Births in last five years | One birth | 2564 | 53.6 |
|  | Two births | 1995 | 41.7 |
|  | More than 2 births | 226 | 4.7 |
| Pregnancy Intention | Unintended | 726 | 15.2 |
|  | Intended | 4059 | 84.8 |
| ANC Received | No | 1324 | 27.7 |
|  | Yes | 3461 | 72.3 |
| Employment Status | Not working | 3124 | 65.3 |
|  | Working | 1661 | 34.7 |
| Age Group | 15-19 | 342 | 7.1 |
|  | 20-24 | 1186 | 24.8 |
|  | 25-29 | 1162 | 24.3 |
|  | 30-34 | 931 | 19.5 |
|  | 35-39 | 779 | 16.3 |
|  | 40-44 | 313 | 6.5 |
|  | 45-49 | 72 | 1.5 |
| Media Exposure | No exposure | 788 | 16.5 |
|  | Low exposure | 2957 | 61.8 |
|  | High exposure | 1040 | 21.7 |
| Women Autonomy | No | 2147 | 44.9 |
|  | Yes | 2638 | 55.1 |

On pregnancy intentions, 84.9% of pregnancies were intended, and 15.2% were unintended, showing high rates of planned pregnancies. Antenatal care engagement is positive, with 72.3% attending at least one visit, while 27.7% did not. Employment data shows 65.3% are not working, and 34.7% are employed, indicating a high rate of unemployment. The age distribution is youthful, with the largest groups being 20-24 years (24.8%) and 25-29 years (24.3%), and only 1.5% aged 45-49. Media exposure varies, 61.8% have low exposure, 16.5% no exposure, and 21.7% high exposure, indicating differing access levels. Women’s autonomy data reveals that 55.1% of respondents have decision making ability, while 44.9% do not.

### Determinants of ANC in Urban and Rural Residences

Table 2 presents the prevalence of receiving antenatal care in relation to various background characteristics and highlights the significance of these associations in both urban and rural setting of Senegal. Region (p<0.05) and age category of respondents (p<0.05) significantly influences the likelihood of antenatal care and it is observed from the table that the percentage of receiving ANC is higher for most urban regions compared to that of rural regions. Moreover, respondent’s education level is also statistically significant (p=0.000) with the association of receiving ANC and respondents with higher education level had the highest percentage of receiving ANC in both residences (Urban = 94.4%, Rural = 92.6%). Wealth index, total number of children born and birth in last five years of respondent also significantly influence the likelihood of receiving ANC (p<0.05) and rich people category had the highest percentage of receiving ANC compared to the remaining groups (Urban = 83.9%, Rural = 76.8%). Moreover, respondents who had less than two children and experienced only one birth in last five years were observed to have highest ANC receiving percentage.

**Table 2:** Bivariate Association of Covariates and ANC in Urban and Rural Residences.

| Variable | Category | Urban (%) |  | Rural (%) |  |
| --- | --- | --- | --- | --- | --- |
|  |  | ANC Received | p value | ANC Received | p value |
| Region | Sedhiou | 72.2 | <0.001 | 47.0 | <0.001 |
|  | Dakar | 85.3 |  | 76.5 |  |
|  | Ziguinchor | 87.5 |  | 64.3 |  |
|  | Diourbel | 87.3 |  | 69.9 |  |
|  | Saint-Louis | 73.5 |  | 62.3 |  |
|  | Tambacounda | 80.3 |  | 76.5 |  |
|  | Kaolack | 84.2 |  | 79.6 |  |
|  | Thiès | 75.9 |  | 71.3 |  |
|  | Louga | 86.3 |  | 80.6 |  |
|  | Fatick | 74.7 |  | 75.8 |  |
|  | Kolda | 81.4 |  | 67.2 |  |
|  | Matam | 70.0 |  | 64.3 |  |
|  | Kaffrine | 88.7 |  | 69.7 |  |
|  | Kedougou | 66.3 |  | 57.1 |  |
| Age Category | 15-19 | 82.1 | <0.001 | 70.8 | 0.001 |
|  | 20-24 | 83.7 |  | 71.9 |  |
|  | 25-29 | 82.1 |  | 69.6 |  |
|  | 30-34 | 76.0 |  | 67.0 |  |
|  | 35-39 | 80.4 |  | 61.7 |  |
|  | 40-44 | 73.0 |  | 71.2 |  |
|  | 45-49 | 58.3 |  | 52.1 |  |
| Education Category | No education | 75.2 | <0.001 | 64.6 | <0.001 |
|  | Primary | 80.3 |  | 73.5 |  |
|  | Secondary | 83.1 |  | 77.7 |  |
|  | Higher | 94.4 |  | 92.6 |  |
| Wealth Index | Poor | 71.4 | <0.001 | 66.4 | <0.001 |
|  | Middle | 77.7 |  | 75.3 |  |
|  | Rich | 83.9 |  | 76.8 |  |
| Total Born Children | Less than 2 | 85.2 | <0.001 | 74.4 | <0.001 |
|  | 2 to 4 | 81.2 |  | 70.1 |  |
|  | More than 4 | 70.3 |  | 62.4 |  |
| Births in last five years | One birth | 83.0 | 0.001 | 71.9 | <0.001 |
|  | Two births | 75.4 |  | 65.8 |  |
|  | More than 2 births | 75.4 |  | 56.8 |  |
| Pregnancy intention | Unintended | 74.5 | 0.018 | 64.0 | 0.029 |
|  | Intended | 80.9 |  | 69.1 |  |
| Media exposure | No exposure | 63.6 | <0.001 | 58.1 | <0.001 |
|  | Low exposure | 80.3 |  | 70.9 |  |
|  | High exposure | 82.4 |  | 72.4 |  |
| Respondent Occupation | Not employed | 79.6 | 0.734 | 69.4 | 0.052 |
|  | Employed | 80.3 |  | 65.9 |  |
| Women autonomy | No | 77.7 | 0.093 | 64.9 | <0.001 |
|  | Yes | 81.1 |  | 71.8 |  |

Intended pregnancies have higher ANC receiving percentage among urban (80.9%) and rural (69.1%) residences and the relationship is significant in both cases. Media exposure is also significantly associated with ANC and high exposure of media resulted in higher ANC receiving percentage. Respondent occupation is not significantly associated with ANC in urban residences but marginally significant in rural residences. Additionally, women autonomy is statistically significant with ANC in rural residences and women with autonomy has higher ANC receiving percentage. On the contrary, women autonomy is not a significant factor for ANC in urban residences.

Table 3 illustrates the findings from the multiple logistic regression analysis, examining the relationship between various categories and ANC attendance compared to their reference groups in urban and rural residences in Senegal. Compared to Sedhiou, urban respondents residing in Ziguinchor, Diourbel, Kaolack and Kaffrine and rural respondents residing in Diourbel, Tambacounda, Kaolack, Thies, Louga, Fatick, Kolda, Matam and Kaffrine had significant higher odds of receiving ANC. Compared to uneducated people, higher educated people are 3.649 times more likely to receive ANC in urban residences and 5.79 times more likely to receive ANC in rural residences and the relationship is significant. Primary and secondary educated people also had higher odds although this relationship is insignificant in urban residences but significant in rural residences.

**Table 3:**
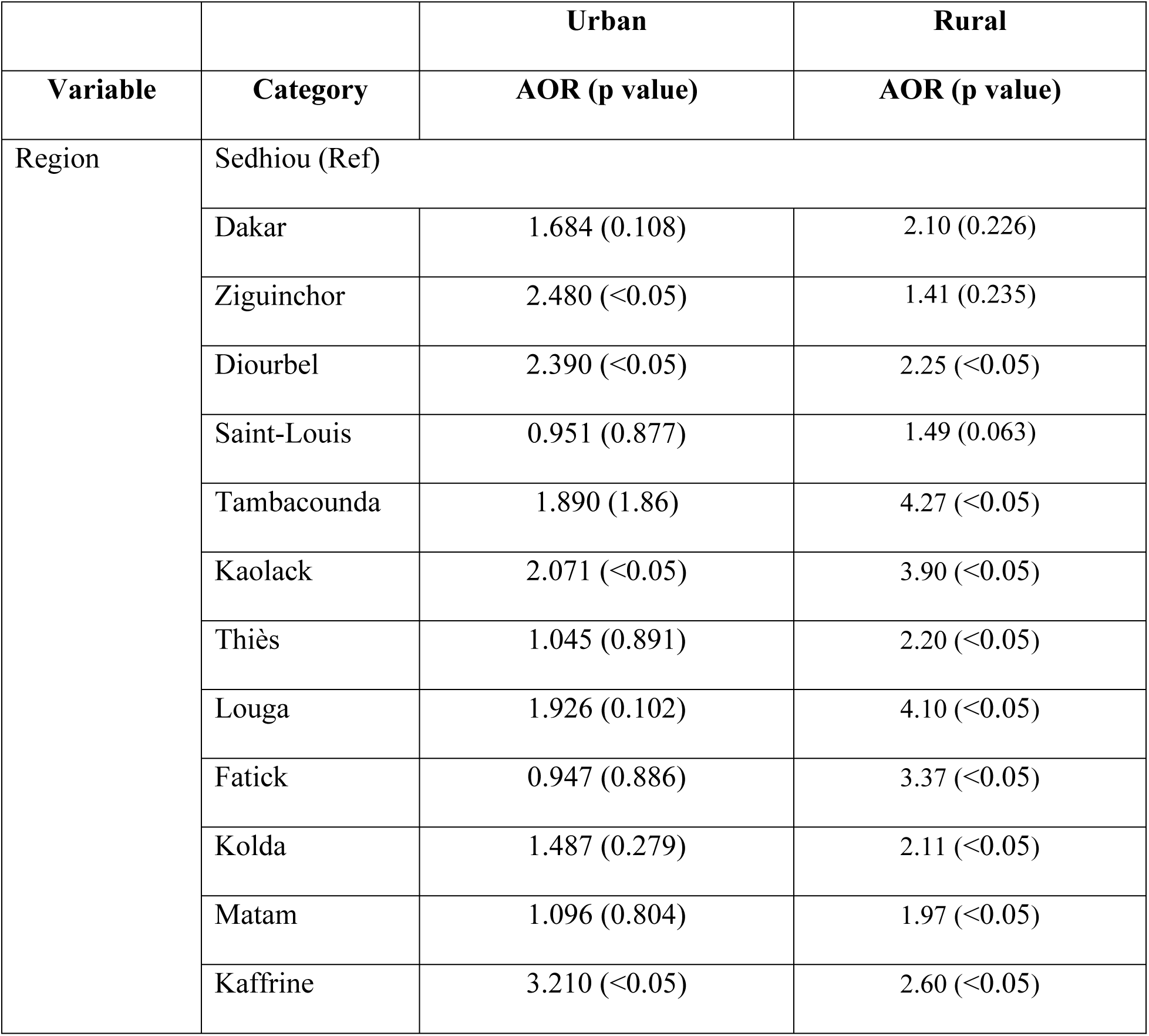
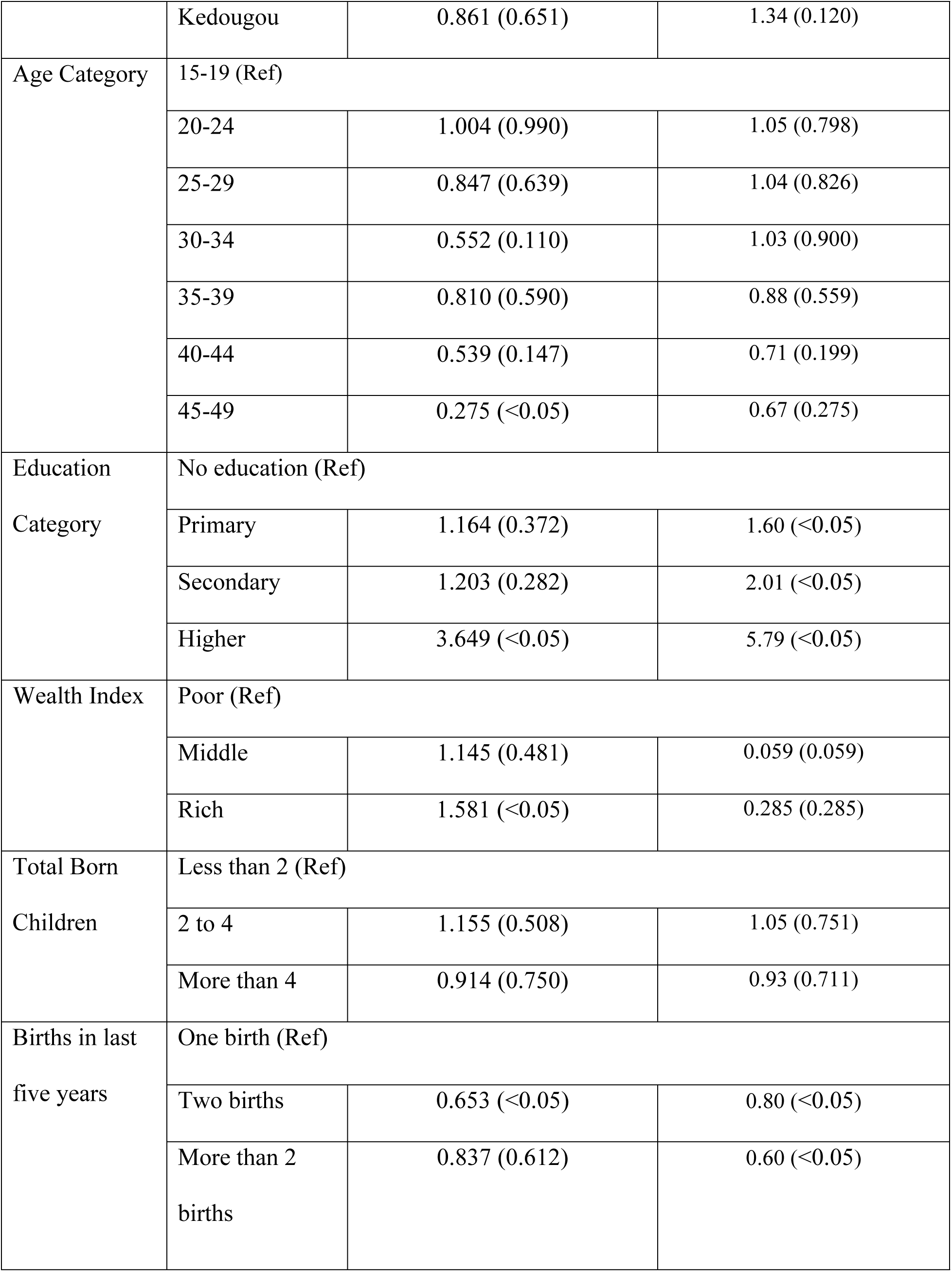
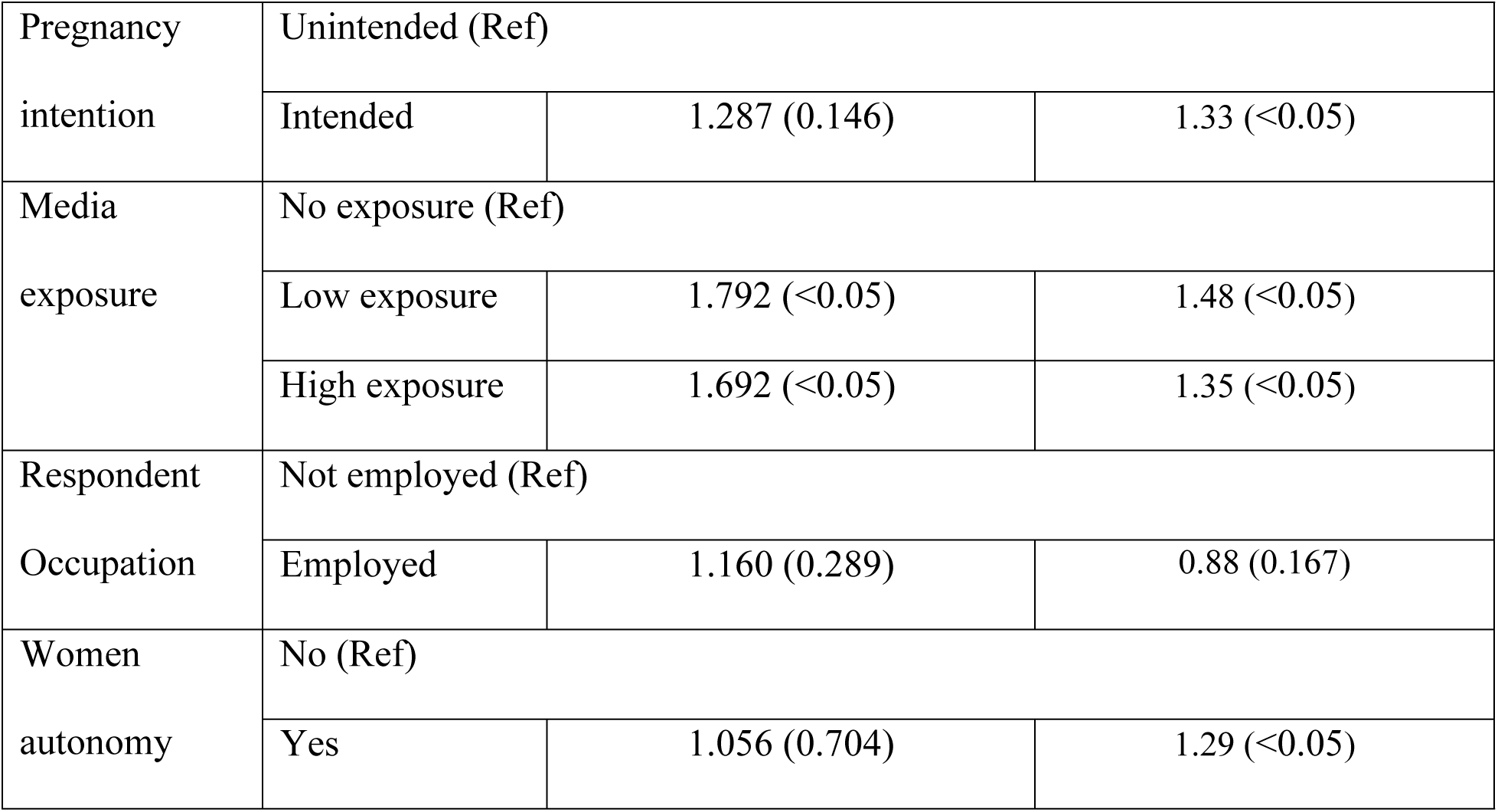
Adjusted Odds Ratio (AOR) with p value obtained from Multiple Logistic Regression.

|  |  | Urban | Rural |
| --- | --- | --- | --- |
| Variable | Category | AOR (p value) | AOR (p value) |
| Region | Sedhiou (Ref) |  |  |
|  | Dakar | 1.684 (0.108) | 2.10 (0.226) |
|  | Ziguinchor | 2.480 (<0.05) | 1.41 (0.235) |
|  | Diourbel | 2.390 (<0.05) | 2.25 (<0.05) |
|  | Saint-Louis | 0.951 (0.877) | 1.49 (0.063) |
|  | Tambacounda | 1.890 (1.86) | 4.27 (<0.05) |
|  | Kaolack | 2.071 (<0.05) | 3.90 (<0.05) |
|  | Thiès | 1.045 (0.891) | 2.20 (<0.05) |
|  | Louga | 1.926 (0.102) | 4.10 (<0.05) |
|  | Fatick | 0.947 (0.886) | 3.37 (<0.05) |
|  | Kolda | 1.487 (0.279) | 2.11 (<0.05) |
|  | Matam | 1.096 (0.804) | 1.97 (<0.05) |
|  | Kaffrine | 3.210 (<0.05) | 2.60 (<0.05) |
|  | Kedougou | 0.861 (0.651) | 1.34 (0.120) |
| Age Category | 15-19 (Ref) |  |  |
|  | 20-24 | 1.004 (0.990) | 1.05 (0.798) |
|  | 25-29 | 0.847 (0.639) | 1.04 (0.826) |
|  | 30-34 | 0.552 (0.110) | 1.03 (0.900) |
|  | 35-39 | 0.810 (0.590) | 0.88 (0.559) |
|  | 40-44 | 0.539 (0.147) | 0.71 (0.199) |
|  | 45-49 | 0.275 (<0.05) | 0.67 (0.275) |
| Education Category | No education (Ref) |  |  |
|  | Primary | 1.164 (0.372) | 1.60 (<0.05) |
|  | Secondary | 1.203 (0.282) | 2.01 (<0.05) |
|  | Higher | 3.649 (<0.05) | 5.79 (<0.05) |
| Wealth Index | Poor (Ref) |  |  |
|  | Middle | 1.145 (0.481) | 0.059 (0.059) |
|  | Rich | 1.581 (<0.05) | 0.285 (0.285) |
| Total Born Children | Less than 2 (Ref) |  |  |
|  | 2 to 4 | 1.155 (0.508) | 1.05 (0.751) |
|  | More than 4 | 0.914 (0.750) | 0.93 (0.711) |
| Births in last five years | One birth (Ref) |  |  |
|  | Two births | 0.653 (<0.05) | 0.80 (<0.05) |
|  | More than 2 births | 0.837 (0.612) | 0.60 (<0.05) |
| Pregnancy intention | Unintended (Ref) |  |  |
|  | Intended | 1.287 (0.146) | 1.33 (<0.05) |
| Media exposure | No exposure (Ref) |  |  |
|  | Low exposure | 1.792 (<0.05) | 1.48 (<0.05) |
|  | High exposure | 1.692 (<0.05) | 1.35 (<0.05) |
| Respondent Occupation | Not employed (Ref) |  |  |
|  | Employed | 1.160 (0.289) | 0.88 (0.167) |
| Women autonomy | No (Ref) |  |  |
|  | Yes | 1.056 (0.704) | 1.29 (<0.05) |

According to wealth index, middle and rich urban people have higher odds of receiving ANC, 1.145 and 1.58 respectively and the relationship is significant for rich people only. On the other hand, rural middle and rich people are less likely to receive ANC compared to that of poor people and this relationship is insignificant. Parity had no significant relationship with ANC among urban and rural respondents. Pregnancy intention resulted in higher odds of receiving ANC, 1.287 and 1.33 odds ratio respectively among urban and rural respondents compared to unintended pregnancy and this relationship is only significant among rural respondents. On the contrary, media exposure became significantly associated with ANC in both residences and exposure to media resulted in higher odds of receiving ANC compared to no exposure to media. Respondent occupation had no significant association with ANC in both residences. However, women autonomy was significantly associated with ANC among rural women but not in urban women and respondents having decision making ability in rural residences were 1.29 times more likely to receive ANC in Senegal.

## Discussion

The objective of the study was to identify different factors associated with the utilization of a minimum of 4 antenatal care services in urban and rural residences in Senegal. This research was conducted based on the nationally representative 2023 Senegal Demographic and Health Survey data where a total of 4785 urban and rural respondents were included who had minimum one birth in last five years prior to the survey. Among the respondents, 72.3% have utilized a minimum of four ANC services in Senegal.

Our study findings reveal that, compared to Sedhiou, women from Ziguichor, Diourbel, Kaolack and Kaffrine in urban residences and women from Diourbel, Tambacounda, Kaolack, Thies, Louga, Fatick, Kolda, Matam and Kaffrine had higher likelihood of receiving in rural residence had higher likelihood of receiving ANC and this relationship is significant. Higher age also resulted in reduced ANC seeking behavior in both urban and rural residence in Senegal which is supported by a similar study in Ethiopia [10]. Moreover, women’s education was positively associated with ANC utilization in Senegal. As the level of education increased from primary to higher, women were more likely to receive ANC in both urban and rural residences compared to uneducated women. This result aligned with a similar study conducted in Nigeria were education was one of the significant factors of receiving ANC [11]. However, our study revealed that all education level had significant association with ANC in rural residences whereas only higher education level was significantly associated with ANC in urban residences in Senegal.

According to wealth index, women from middle and rich households are likely to have more ANC in urban residences and this relationship is significant only rich household but the likelihood decreases in rural residences and none of the wealth index levels are significant with ANC for rural residences compared to poor households in Senegal. This outcome is in line with a similar study conducted in Ghana where wealth indexes in rural residences were insignificant although it was significant for urban residences [12]. Moreover, respondents who had experienced more than one births in last five years in both urban and rural residences were less likely to receive ANC compared to those who experienced only one child birth and this relationship is significant in both urban and rural residences in Senegal. This outcome aligned with a similar study conducted in Ghana where it was revealed that more than four births had a decreasing likelihood of ANC [13].

Intended pregnancies had a higher likelihood of receiving ANC compared to unintended pregnancies in both residences though this relationship is only significant among rural women. A systematic review study identified that unintended pregnancy resulted in delayed antenatal care compared to intended pregnancy and our results also agrees with it [14]. Media exposure is an important factor for ANC and it is revealed that respondents who were exposed to some media had a higher likelihood of receiving ANC in both residences. Several similar studies have also identified media exposure as an important factor for receiving ANC [15-16]. However, the likelihood is slightly higher in urban residences and this relationship is significant among both urban and rural women in Senegal. Respondent who are employed displayed a higher likelihood of receiving ANC among urban women but lower likelihood among rural women. However, this relationship is insignificant in both residences in Senegal. Our study did not agree to a similar study conducted in East African countries where respondent’s occupation had a significant impact on receiving ANC [17]. Women who participated in household decision making resulted in increased likelihood of ANC in both urban and rural residences. This outcome aligned with the results in India where women autonomy resulted in increased likelihood of receiving ANC [18]. However, this relationship is not significant in urban residences but significant in rural residences.

## Limitations of the Study

The study was conducted on Senegal DHS which relies on self-reported information. Hence the data can be subject to recall bias or misreporting. Additionally, the study utilized data obtained from a cross sectional survey, which can only demonstrate the presence of an association between variables, rather than a cause-and-effect relationship.

## Conclusion

The study revealed a noticeable difference among the factors responsible for receiving ANC in urban and rural residences of Senegal. Higher level of education was a significant factor of ANC in both residences although, only in rural residences, primary and secondary education also had a significant influence. This finding underscores the importance of enhancing educational opportunities in rural settings as a critical policy lever for improving maternal health outcomes. On the contrary, wealth index was significantly associated with ANC in urban respondents but it had no impact on rural area women. Women exposed to media are more likely to receive ANC in both areas and this relationship is significant and so raising awareness via various media play a crucial role of receiving ANC in both urban and rural residences. Women autonomy only has significant influence on ANC in rural areas but not in urban residences which highlights the role of woman empowerment in rural regions for receiving ANC. The different factors identified as determinants of attending four or more ANC visits among urban and rural respondents in this study offer valuable insights to adjust its policy initiatives aimed at boosting the coverage of four or more ANC visits among women in Senegal.

## Acknowledgements

The authors thank Prof. Dr. Wasimul Bari for his useful comments and suggestions.

## Authors’ contributions

MRKM, SAT and GK generated the study idea, formed the research team, and designed the study and conducted the initial literature review. SAT wrote the introduction, MRKM, SAT and GK wrote the methodology and MRKM analyzed the data. MRKM wrote the initial draft of the manuscript. All authors contributed to the intellectual content of the manuscript and read and approved the final version of the manuscript.

## Ethics approval and consent to participate

As the data was obtained from an open source of Official Website of DHS (https://dhsprogram.com/), the ethical approval and consent to participate are not applicable for this study.

## Consent for publication

As the study does not contain any individual and institutional data in any form (including any individual details, images, or videos), the consent for publication is not applicable for this study.

## Availability of data and materials

The dataset, analyzed in this study, was obtained from the open source of Official Website of DHS program (Weblink: https://www.dhsprogram.com/Data/). The Senegal Demographic and Health Survey 2023 dataset was availed from website for this study (Link: https://dhsprogram.com/methodology/survey/survey-display-611.cfm)

